# Glycaemic outcomes of three months of continuous glucose monitoring in children, adolescents and young adults with type 1 diabetes at a Kenyan tertiary hospital: a single-arm prospective study

**DOI:** 10.64898/2026.09.29.26364323

**Authors:** Lucy Wainaina Mungai, Prisca Amolo, Agnes Karingo Karume, Purity Munene-Waweru, Catherine Nailaen, Philip Mule, Nuria Botella, Priyanka Singh, Elena Marbán-Castro, Yvonne Kamau

**Author notes:** Corresponding author: Lucy Mungai, Department of Paediatrics and Child Health, University of Nairobi, PO Box 19676-00202, Nairobi, Kenya.

## Abstract

Continuous glucose monitoring (CGM) is recommended for type 1 diabetes, but the supporting evidence derives almost entirely from high-income settings, and the technology is not routinely available in Kenya’s public sector. This prospective study assessed the feasibility and sustainability of CGM implementation and changes in glycemic control after three months of use in a public-sector pediatric diabetes clinic in Nairobi. Forty patients aged 4 to 25 years, recruited by proportional quota sampling across age bands at Kenyatta National Hospital, used an intermittently scanned sensor system across five visits combining sensor data review, standardized diabetes education and insulin titration. Glycated hemoglobin (HbA1c) was measured at baseline and three months, alongside sensor-derived glucose metrics, wear time and diabetes-related admissions. Thirty-eight patients (95%) completed the study; median sensor wear time was 89.0%. Adjusted mean HbA1c fell from 10.82% to 9.42%, a difference of −1.40 percentage points (95% confidence interval −2.07 to −0.68). Time in target range (3.9 to 10.0 mmol/L) rose from 32.4% to 38.0%, and time above 10.0 mmol/L fell from 63.1% to 56.9%. Time below 3.9 mmol/L rose minimally, from 3.7% to 4.2%, while time below 3.0 mmol/L did not increase significantly, by 0.2%. Diabetes-related admissions fell from 7 of 40 in the three months before the study to 1 of 38 by its end. The uncontrolled design and small sample preclude an unbiased effect estimate. Since CGM efficacy is established internationally, these data add local evidence of transferability, which national guidelines and benefit-package decisions require.

## 1. Introduction

In type 1 diabetes (T1D) of childhood and adolescence, sustained hyperglycaemia drives micro- and macrovascular disease, and complications are greatest with early onset as exposure may span decades [1]. The Diabetes Control and Complications Trial (DCCT) showed that intensive glycaemic management reduces retinopathy, nephropathy and neuropathy risk by 34–76% at a mean glycated haemoglobin (HbA1c) of about 7.0% [1], and in the Epidemiology of Diabetes Interventions and Complications (EDIC) follow-up this protection persisted and extended to cardiovascular outcomes [20].

Though HbA1c is a validated marker of mean glycemic exposure, it describes glucose distribution poorly. It is insensitive to glycemic variability, frequency and duration of hypoglycemia, and postprandial glucose excursions. High glycemic variability, expressed as the coefficient of variation (CV), has been shown to predict hypoglycemia risk and endothelial injury [2, 5].

Continuous Glucose Monitoring (CGM) records interstitial glucose every 1-5 minutes, allowing calculation of standardized consensus metrics: time in range (TIR, 3.9–10.0 mmol/L), time above range (TAR, >10.0 mmol/L) and time below range (TBR, <3.9 mmol/L) [5]. In high-income settings, trials show that CGM, compared with self-monitoring of blood glucose (SMBG), lowers HbA1c by 0.4–1.0%, raises TIR by 10–15 percentage points and reduces hypoglycaemia in adult, adolescent and paediatric T1D populations, with meta-analyses reporting consistent effects across health systems [2, 3, 4, 14].

T1D in sub-Saharan Africa is managed under substantially different conditions. Insulin supply is often intermittent because of procurement failures and cold-chain gaps, diagnosis is frequently delayed so that diabetic ketoacidosis (DKA) is common at presentation, specialist care is concentrated in tertiary urban centres, and limited household income restricts monitoring consumables [9, 11, 12]. Mean HbA1c in regional paediatric cohorts therefore far exceeds international targets, DKA and severe hypoglycaemia admissions are frequent, and complication rates are correspondingly high [6, 9, 10].

Regional evidence indicates that CGM is deployable and may confer glycaemic benefit. McClure Yauch and colleagues described the glycaemic burden of an East African paediatric cohort using CGM, recording a mean TIR of about 37% and sustained hyperglycaemia [6]. Studies from Rwanda and Malawi report that CGM is feasible and describe early improvements in glycaemic metrics and self-management [7, 8]. None of these combined prospective pre- and post-intervention HbA1c with CGM-derived distribution and adherence data.

Kenya has among the highest estimated T1D burdens in sub-Saharan Africa. CGM is not routinely available in the public sector, and paediatric glycemic control falls well below international benchmarks [25]. Prospective, context-specific data are needed to inform national guidelines, health technology assessment and advocacy for subsidized access. The ACCEDE protocol set out three-arm pragmatic, randomized trials of CGM devices in South Africa and Kenya 21]; the randomized comparison was conducted at the South African site, while in Kenya the evaluation took the form of the single-arm three month prospective study reported here.

This study was set out to test whether transferability of benefit demonstrated in well-resourced trial settings survives the conditions of a public clinic where human rather than analogue insulin predominates, supply is intermittent, households are of limited means and routine review was six-weekly. Our objectives were therefore to establish whether CGM could be delivered and sustained in this setting, to characterize the cohort’s glycemic profile, and to describe the glycaemic change that accompanied three months of use. We report change in HbA1c, CGM-derived glycaemic distribution metrics, sensor wear time and diabetes-related hospital admissions.

## 2. Materials and Methods

### 2.1. Study design and reporting

This was a prospective, single arm, unblinded study conducted at the paediatric, adolescent and youth endocrinology clinic of Kenyatta National Hospital (KNH), Nairobi, the largest public hospital in East Africa. It was nested within a broader CGM usability study whose acceptability and feasibility findings, drawn from the same participants, are reported separately [22]. The present manuscript reports only the clinical and glycemic outcomes; there is no overlap of results between the two reports. The study is reported in accordance with the TREND statement for non-randomized evaluations [23].

The study was conducted at the Kenyan site of the ACCEDE programme, whose protocol described three-arm pragmatic randomized controlled trials of CGM devices in Kenya and South Africa [21]. The randomized comparison was undertaken in South Africa; the Kenyan evaluation was the single-arm study reported here. Trial registration: Pharmacy and Poisons Board Expert Committee on Clinical Trials (ECCT/25/08/07).

### 2.2. Participants

Individuals aged 4–25 years with a confirmed diagnosis of T1D and in active follow-up at the clinic were eligible. Exclusion criteria were participant refusal to give consent/assent or inability to go through the CGM-related diabetes education. Participants meeting the eligibility criteria were recruited by quota sampling, with quotas allocated proportionally across age bands so that the enrolled cohort mirrored the age distribution of the clinic population. Enrolment across the reporting categories was 11 children aged 4 to under 10 years, 19 adolescents aged 10 to under 18 years, and 10 young adults aged 18 to 25 years. Written informed consent was obtained from participants aged 18 years and above and from the parent or legal guardian of each participant under 18; children aged 7–17 years also gave assent.

### 2.3. Intervention

The intervention was CGM delivered within a package that also included structured education and systematic insulin titration, and these components cannot be separated in this design. At the first sensor insertion (Visit 2), participants and caregivers received standardized education covering sensor application, wear and the interpretation of glucose trend data. At Visits 3, 4 and 5 (two weeks, six weeks and three months), sensor data were downloaded and reviewed with the participant, time in, above and below range were discussed, insulin doses were adjusted where indicated, and insulin-to-carbohydrate ratio and correction factor were reinforced. At Visit 5 participants were counselled on continuing self-management without CGM and referred for diabetes education. Standard management of T1D at this clinic during the study period was scheduled clinic review every six weeks, so the study schedule added approximately two clinical visits over the three months, together with structured CGM education and systematic insulin titration that are not part of routine review. Participants were recruited one month before the CGM insertion and then scheduled for visit 2. The increase in contact frequency was therefore modest, but it was not absent, and all analyses should be read as evaluating CGM delivered within this package rather than CGM alone.

### 2.4. CGM device and data handling

Participants used an intermittently scanned CGM system (FreeSyle Libre, Abbott Diabetes Care Inc. Model Number: 71992, Second generation), with glucose values visible to the participant on scanning rather than blinded for later review. Sensors were worn continuously for 10–14 days per manufacturer’s specifications. A replacement sensor was provided when a sensor detached prematurely, and detached sensors were returned at the next visit; participants received approximately 7 sensors on average over the three months.

Sensor wear time was defined as the percentage of days across the three-month period on which sensor data were recorded. For the glycemic distribution analyses, a participant’s data for a given biweekly period were considered evaluable when at least 70% of possible readings over 14 days were available, in line with the international consensus on CGM data sufficiency [5, 24]. Data were entered into REDCap (Vanderbilt University, Nashville, TN, USA).

### 2.5. HbA1c measurement

HbA1c was measured on capillary blood by point-of-care assay (Affinion 2, Abbott Diagnostics) at baseline and at three months, using the same instrument and operator procedure at both timepoints. The assay is National Glycohemoglobin Standardization Program (NGSP-) certified and International Federation of Clinical Chemistry and Laboratory Medicine (IFCC)-aligned. The method is based on boronate affinity separation, which is comparatively insensitive to the common hemoglobin variants, including hemoglobin S and C, that are prevalent in this population; nevertheless, HbA1c may underestimate mean glycemia in the presence of anemia or shortened erythrocyte survival, and hemoglobinopathy status was not systematically ascertained in this cohort.

### 2.6. Outcomes

The primary outcome was change in HbA1c from baseline to three months. Secondary outcomes were CGM-derived glycaemic metrics (TIR, TAR, TBR, mean interstitial glucose and CV), sensor wear time, and diabetes-related hospital admissions. Admissions in the three months preceding enrolment were ascertained by participant and caregiver recall at Visit 1, whereas admissions during follow-up were captured prospectively at study visits; the two periods are therefore not ascertained by the same method.

### 2.7. Sample size

The sample size of 40 was determined by the objectives of the parent usability study, a formal power calculation was therefore not undertaken. Because the efficacy of CGM is already established in randomized trials, the purpose here was to establish deliverability and to estimate the glycemic change accompanying use in this setting rather than to re-test efficacy [2, 3, 4, 14]. Consistent with guidance for pilot studies, results are presented as estimates with 95% confidence intervals so that the precision achieved is explicit.

### 2.8. Statistical analysis

Categorical variables are presented as frequencies and percentages and continuous variables as medians with interquartile ranges (IQR), given the non-normal distribution of key clinical parameters. Glycemic variability was quantified as the coefficient of variation (CV = SD/mean × 100), with CV <36% taken as the threshold for acceptable hypoglycemia risk in T1D [5].

Change in HbA1c from baseline to endline was analyzed using a linear mixed-effects model (LMM) with a random intercept per participant, adjusting for age and sex. Adjusted mean differences (baseline vs. endline) and their corresponding 95% confidence intervals were derived from estimated marginal means (EMMs) using the emmeans package. CGM-derived metrics (TIR, TAR, TBR, CV and interstitial glucose) were summarized as biweekly medians with IQRs and presented graphically.

### 2.9. Ethic approval

The study complied with the Declaration of Helsinki. Ethical approval was granted by the Kenyatta National Hospital–University of Nairobi Ethics and Research Committee (P291/03/2025), the Pharmacy and Poisons Board Expert Committee on Clinical Trials approved the study (ECCT/25/08/07), and a research permit was issued by the National Commission for Science, Technology and Innovation (P/2025/4179964).

## 3. Results

### 3.1. Participants and retention

Between November 2025 and January 2026, 40 individuals with T1D were enrolled. Two participants withdrew before the endline assessment: one because the caregiver took up employment away from home and the participant was unable to manage the sensor independently, with multiple sensor detachments; the other because the participant relocated outside the catchment area. Thirty-eight participants (95%) contributed to the primary endline.

Sensor wear time, defined as the percentage of days with recorded sensor data, had a mean of 81.3% (SD 17.6) and a median of 89.0% (IQR 75.0–92.0), with a range of 15.0–95.0%. Eighty percent (32/40) of participants wore the sensor for at least 70% of the period.

### 3.2. Baseline characteristics

Baseline characteristics are shown in Table 1. Median age was 13.0 years (IQR 9.3–17.5), most participants were female (25/40; 62.5%), and adolescents aged 10 to under 18 years formed the largest group (19/40; 47.5%). Median diabetes duration was 5.0 years (IQR 3.0–6.5) and enrolment HbA1c ranged from 6.50% to 15.30%.

**Table 1.** Baseline sociodemographic and clinical characteristics of participants (N = 40).

| **Characteristic** | **Population (N = 40)** |
| --- | --- |
| Sex: female | 25 (62.5%) |
| Age, years, median (IQR) | 13.0 (9.3–17.5) |
| **Age group** |  |
| Children (4 to <10 years) | 11 (27.5%) |
| Adolescents (10 to <18 years) | 19 (47.5%) |
| Young adults (≥18 years) | 10 (25.0%) |
| **Highest level of education** |  |
| Primary | 25 (62.5%) |
| Secondary/A-level | 10 (25.0%) |
| College/middle level | 1 (2.5%) |
| University | 4 (10.0%) |
| **Employment status** |  |
| Employed full-time | 2 (5.0%) |
| Employed part-time | 1 (2.5%) |
| Student | 3 (7.5%) |
| Not in school and unemployed | 4 (10.0%) |
| Not applicable (children/adolescents) | 30 (75.0%) |
| HbA1c at enrolment (%), (IQR) | 10.82 (8.95–11.95) |
| Duration of T1D, years, median (IQR) | 5.0 (3.0–6.5) |
| **SMBG checks per day** |  |

| Characteristic | Population (N = 40) |
| --- | --- |
| 1–3 | 21 (52.5%) |
| 4–6 | 19 (47.5%) |
| **Self-reported hypoglycemic episodes per week** |  |
| 0 | 8 (20.0%) |
| 1–2 | 19 (47.5%) |
| >2 | 13 (32.5%) |
| **DKA episodes in preceding 12 months** |  |
| 0 | 23 (57.5%) |
| 1 | 10 (25.0%) |
| >1 | 7 (17.5%) |
| **Insulin regimen** |  |
| Rapid-acting + long-acting | 7 (17.5%) |
| Short-acting + long-acting | 33 (82.5%) |
| Prior CGM exposure | 4 (10.0%) |

DKA, diabetic ketoacidosis; IQR, interquartile range; SMBG, self-monitoring of blood glucose; T1D, type 1 diabetes.

Just over half of participants (52.5%) tested blood glucose one to three times daily and 47.5% four to six times. Self-recognized hypoglycemia was common: 47.5% reported one to two episodes per week and 32.5% more than two. DKA was frequent, with 42.5% reporting at least one episode in the preceding 12 months and 17.5% more than one. Most participants (33/40; 82.5%) used short-acting with long-acting insulin and only 17.5% (7/40) used rapid-acting analogues with long-acting insulin. Four participants (10%) had prior CGM exposure.

### 3.3. Caregiver characteristics

Caregiver data were available for the 30 participants aged under 18 years (Table 2). Most caregivers were biological parents (28/30; 93%) and female (22/30; 73%), with a median age of 38 years (IQR 32–42). All had attended school, and 95% reported a source of income, although few held full-time employment. Reported household income placed most families in the lower economic strata.

**Table 2.**
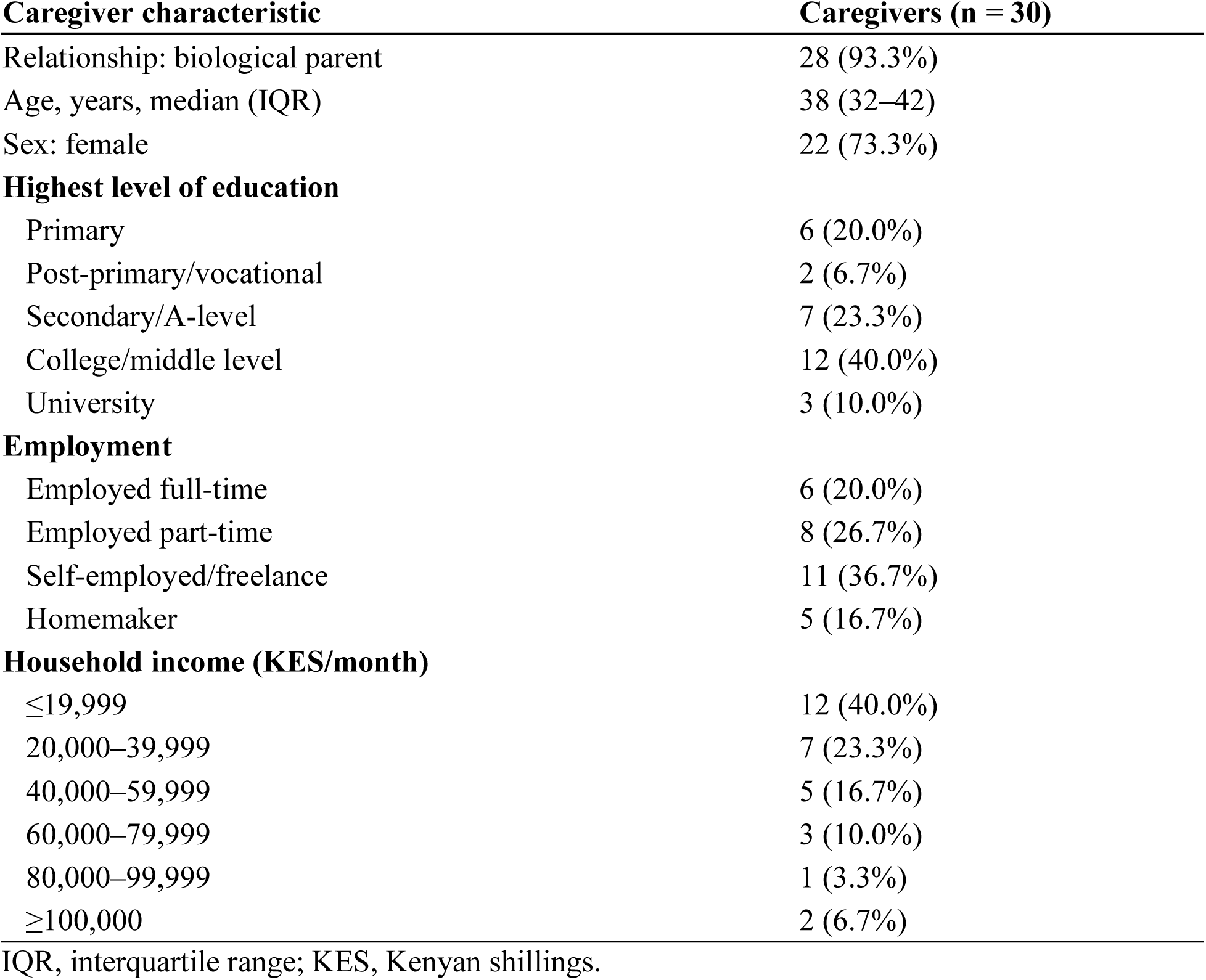
Caregiver characteristics for participants aged under 18 years (n = 30).

| Caregiver characteristic | Caregivers (n = 30) |
| --- | --- |
| Relationship: biological parent | 28 (93.3%) |
| Age, years, median (IQR) | 38 (32–42) |
| Sex: female | 22 (73.3%) |
| **Highest level of education** |  |
| Primary | 6 (20.0%) |
| Post-primary/vocational | 2 (6.7%) |
| Secondary/A-level | 7 (23.3%) |
| College/middle level | 12 (40.0%) |
| University | 3 (10.0%) |
| **Employment** |  |
| Employed full-time | 6 (20.0%) |
| Employed part-time | 8 (26.7%) |
| Self-employed/freelance | 11 (36.7%) |
| Homemaker | 5 (16.7%) |
| **Household income (KES/month)** |  |
| ≤19,999 | 12 (40.0%) |
| 20,000–39,999 | 7 (23.3%) |
| 40,000–59,999 | 5 (16.7%) |
| 60,000–79,999 | 3 (10.0%) |
| 80,000–99,999 | 1 (3.3%) |
| ≥100,000 | 2 (6.7%) |

IQR, interquartile range; KES, Kenyan shillings.

### 3.4. Change in HbA1c

Adjusted mean HbA1c fell from 10.82% at baseline to 9.42% at three months, a difference of −1.40 percentage points (95% CI −2.07 to −0.68; p < 0.001) (Figure 1).

**Figure 1.**
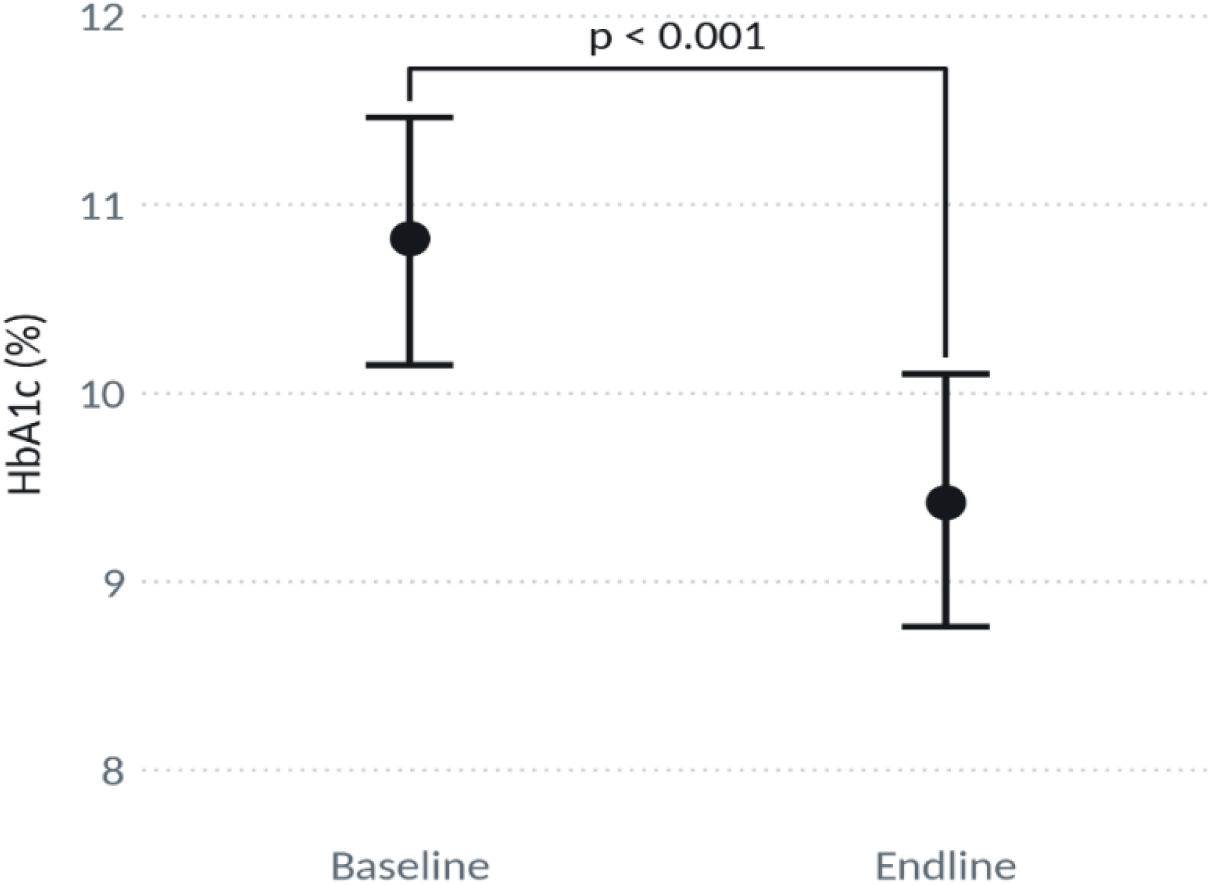
Fitted HbA1c (%) by timepoint.

Points represent the sample mean at each timepoint and vertical bars the 95% confidence interval for the mean obtained from the linear mixed-effects model. The unadjusted paired difference was −1.4 percentage points, indicating that adjustment for age and sex did not materially alter the estimate.

### 3.5. CGM-derived glycemic metrics

CGM-derived metrics at baseline and at three months are presented according to the international consensus definitions, which set the target range at 3.9–10.0 mmol/L, and are shown in Figures 2 to 4 [5].CGM data analyses below cover four metrics: mean interstitial glucose, glycemic variability measured by CV, time in glycemic ranges, and glucose distribution across five clinical categories. All are presented according to the ATTD consensus definitions, which set the target range at 3.9–10.0 mmol/L [5].

**Figure 2.**
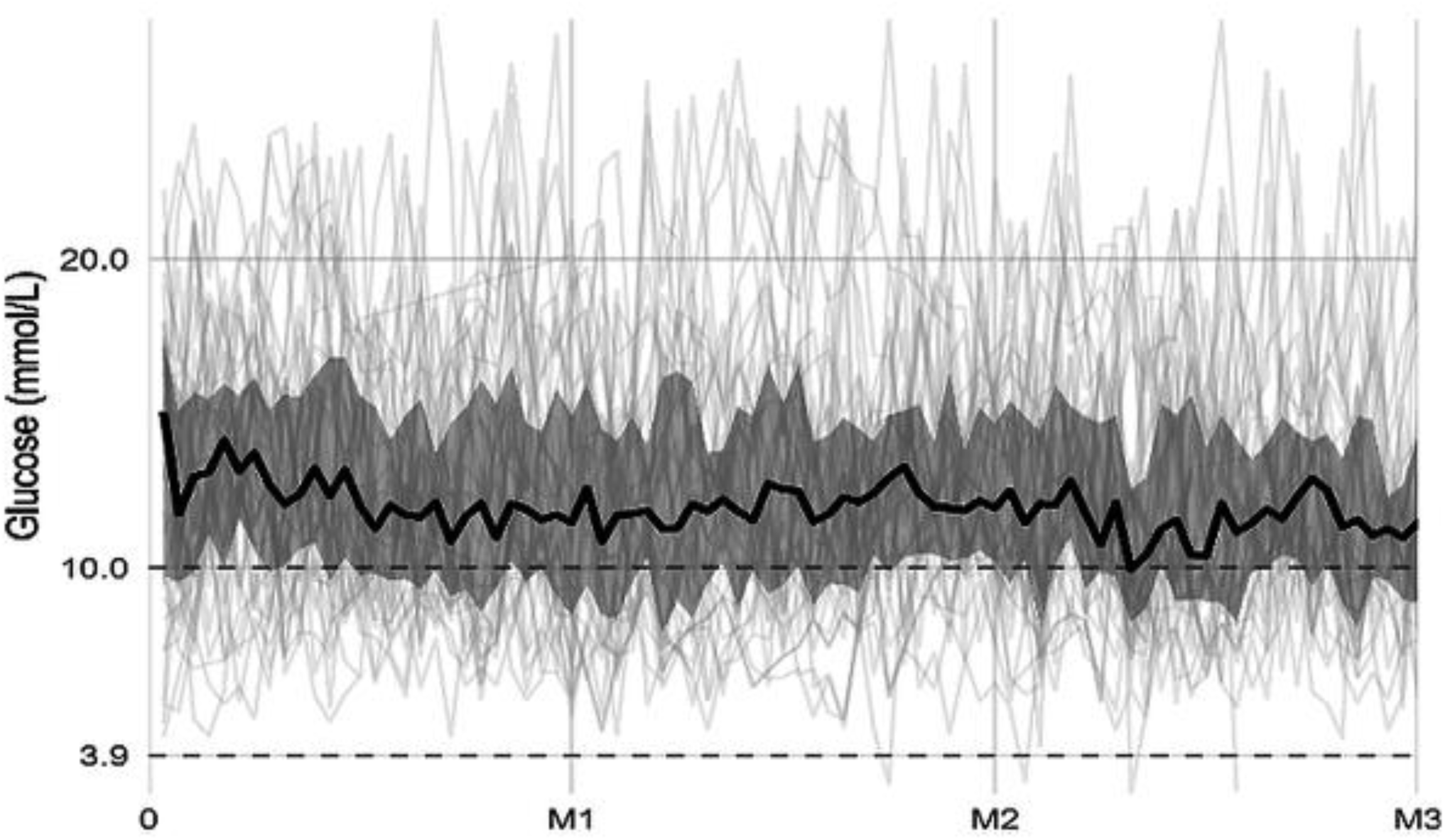
Daily mean interstitial glucose (mmol/L). Line plot with shaded IQR ribbon; solid line, population median; grey lines, individual trajectories; dashed lines at 3.9 and 10.0 mmol/L marking the target range.

#### 3.5.1. Mean interstitial glucose

Figure 2 shows the daily mean interstitial glucose over the three months and the interquartile range used by participants to incorporate CGM trend data into insulin decisions. Mean interstitial glucose fell from 15.0 mmol/L at baseline to 11.5 mmol/L at three months, a reduction of 3.5 mmol/L. Variation between individuals was wide at baseline, matching the range of enrolment HbA1c values (6.50–15.30%). By the later weeks, a substantial proportion of individual trajectories had moved towards or into the target range of 3.9–10.0 mmol/L, aligned with the HbA1c reduction reported above.

#### 3.5.2. Glycemic variability (coefficient of variation)

Glycemic variability, expressed as the coefficient of variation, is shown in Figure 3.

**Figure 3.**
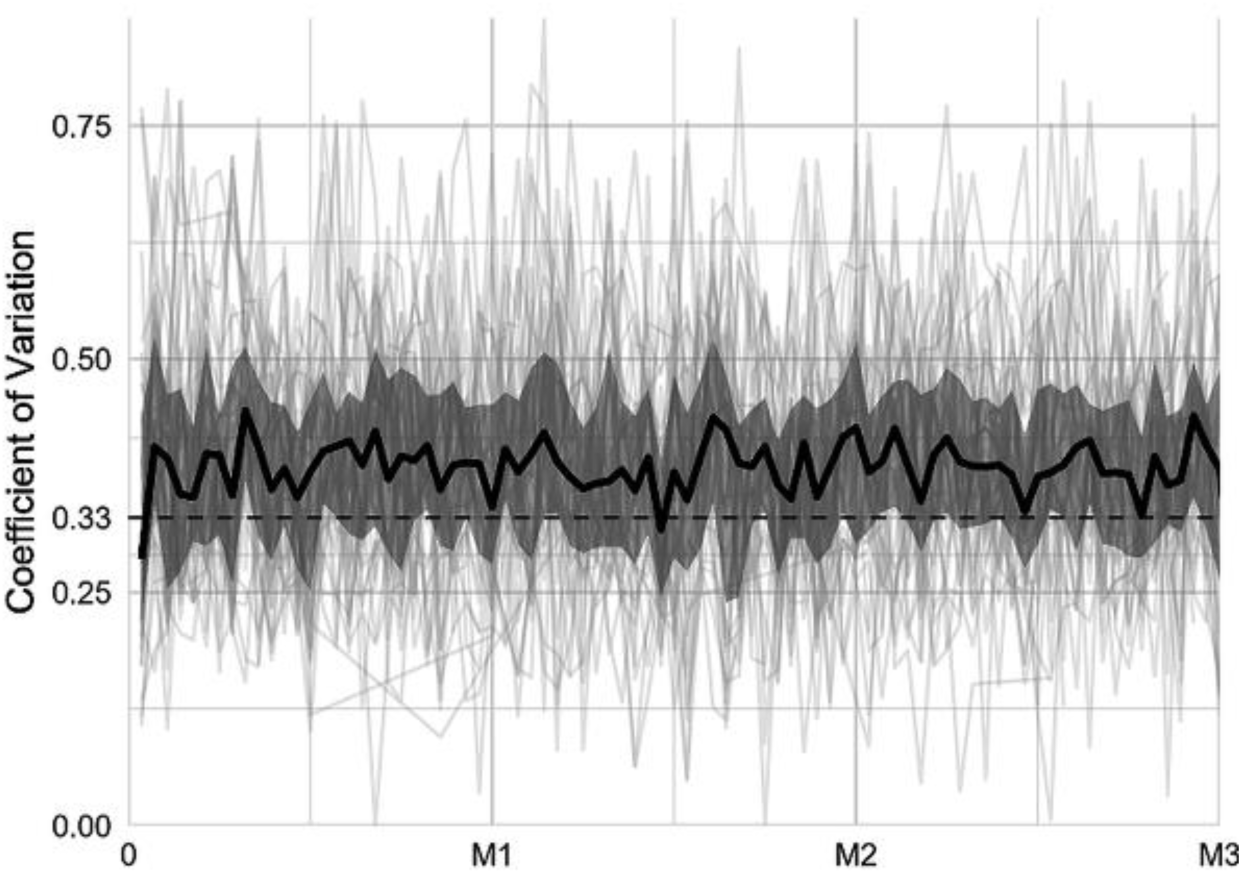
Coefficient of variation of interstitial glucose. Solid line, population median; shaded ribbon, IQR; dashed line at the 36% threshold.

The population median tracked close to the 36% threshold throughout the three months, with wide dispersion between individuals at every timepoint.

The variability therefore did not increase as mean glucose fell, indicating that the reduction in hyperglycemia reported above was not achieved at the cost of greater instability.

#### 3.5.3. Glycemic distribution

TIR (3.9–10.0 mmol/L) rose from 32.4% to 38.0%, an increase of 5.6 percentage points, and time above 13.9 mmol/L fell from 43.2% to 36.1%, a reduction of 7.1 percentage points, while time between 10.0 and 13.9 mmol/L was essentially unchanged (19.9% to 20.8%). TBR rose by 0.5% in the low band and stayed consistently low in the very low band.

Overall, TIR rose across successive biweekly periods, with a proportionate fall in TAR (Figure 4). This matches the downward trend in mean glucose and shows how values were redistributed across the clinical range.

**Figure 4.**
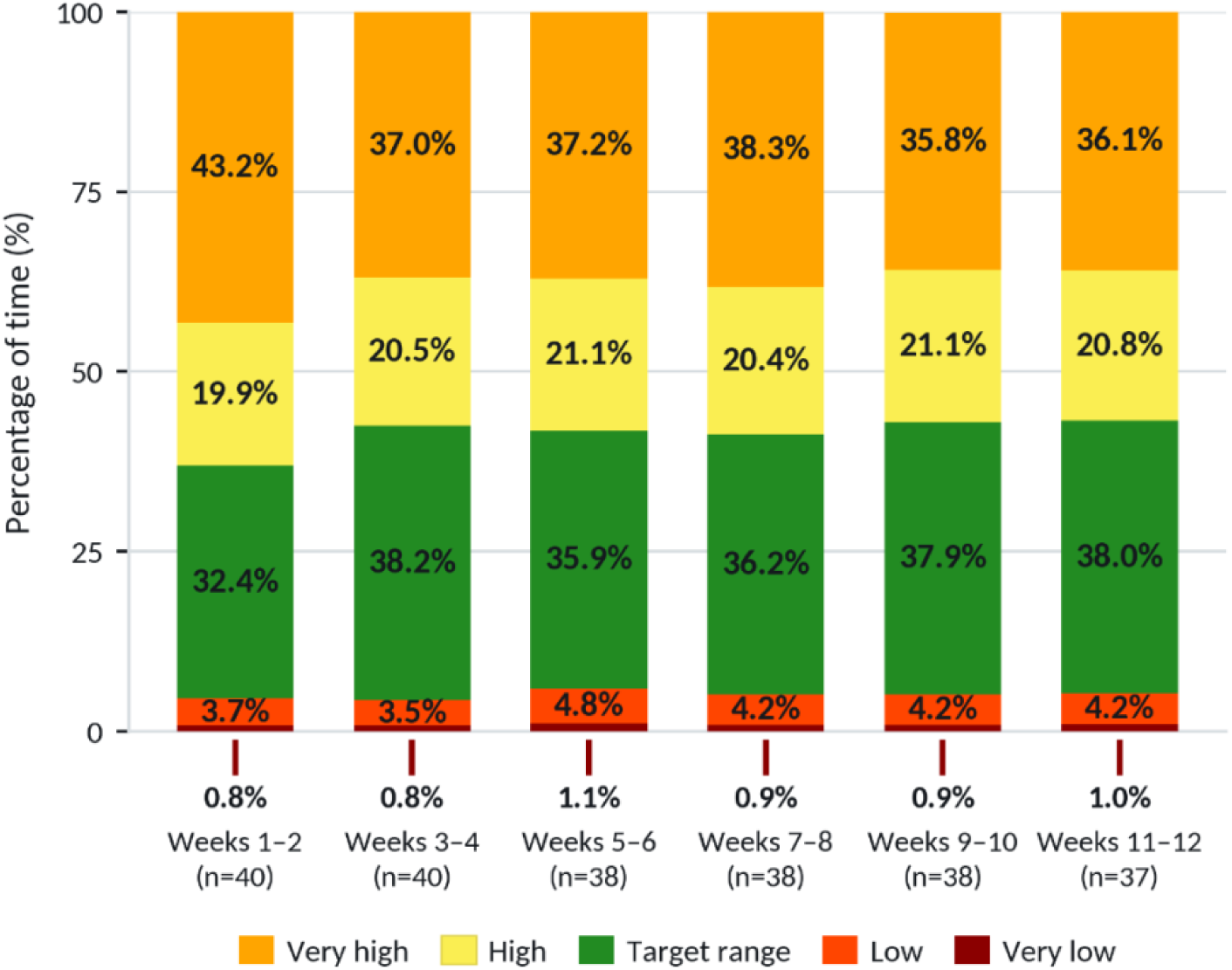
Biweekly glycemic distribution across five clinical categories: very high >13.9; high 10.1–13.9; target 3.9–10.0; low 3.0–3.8; very low <3.0 mmol/L.

The proportion of values above 13.9 mmol/L showed the steepest proportional decline; the most dangerous extreme of the distribution improved most under CGM-informed management. The low (3.0–3.8 mmol/L) and very low (<3.0 mmol/L) categories remained small throughout and showed no clinically meaningful increase.

### 3.6. Hospital admissions

Diabetes-related admissions fell from 7/40 (17.5%) in the three months before enrolment to 1/38 (2.6%) during the three months of CGM use (Table 3). The reasons differed: six of the seven earlier admissions were for DKA and one was a hyperglycemia-related presentation (cystopathy with poorly controlled T1D), whereas the single admission during the study was for hypoglycemia. These proportions are presented descriptively; the numbers are too small to support inference, and the pre-enrolment period was ascertained by recall while the study period was captured prospectively.

**Table 3.**
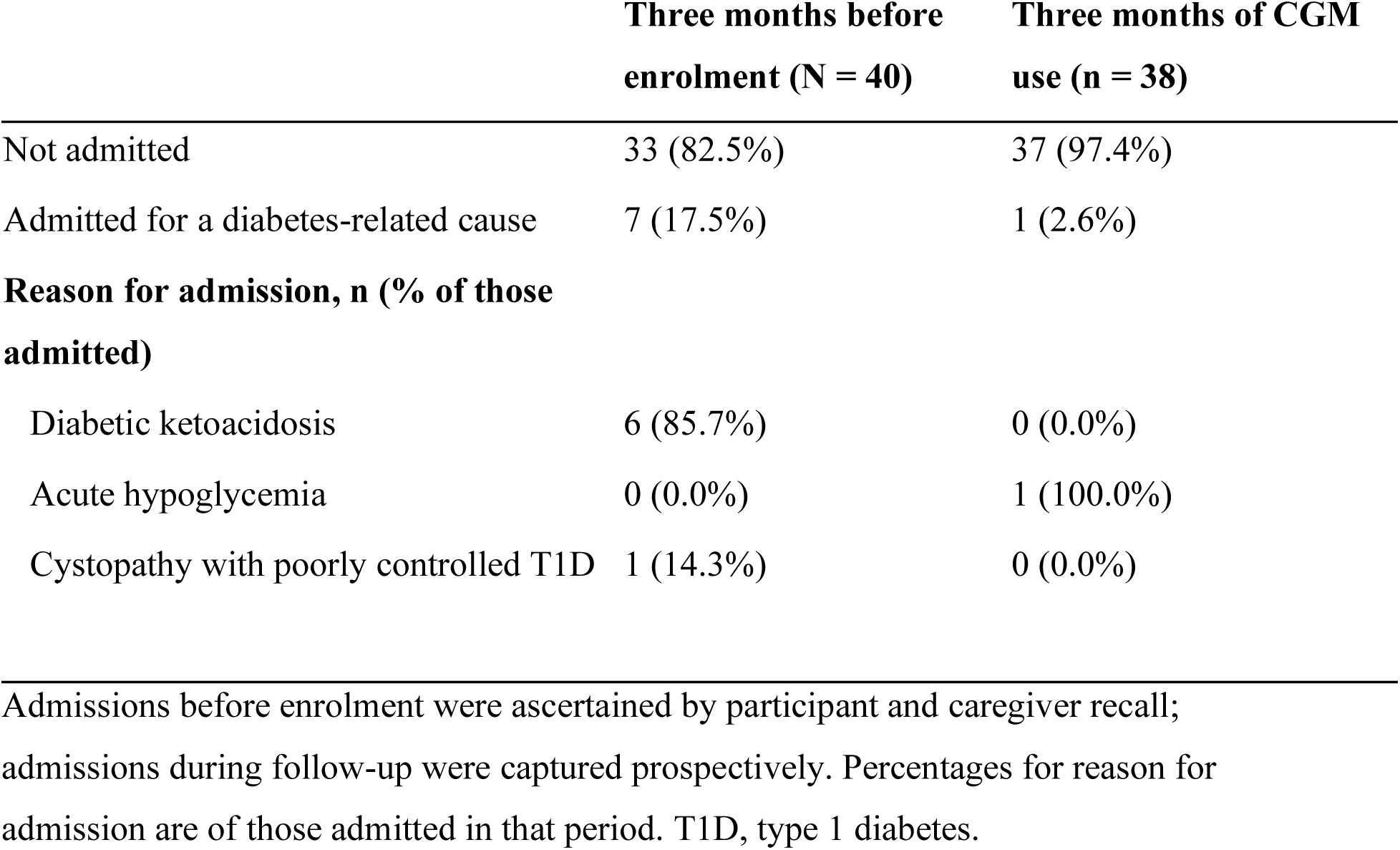
Diabetes-related hospital admissions in the three months before enrolment and during the three months of continuous glucose monitoring use.

## 4. Discussion

This study is one of the few from sub-Saharan Africa that gives prospective CGM data. In this single-arm pilot at a national referral hospital in Kenya, three months of CGM delivered alongside structured education and systematic insulin titration was associated with a fall in adjusted mean HbA1c from 10.82% to 9.42%, with high sensor adherence and without a clinically meaningful increase in time spent below 3.0 mmol/L. CGM-derived metrics moved consistently, with TAR falling across successive biweekly periods and TIR rising, though remaining below the recommended 70% target [5]. The low burden of severe hypoglycemia matters in a setting where glucagon rescue is rarely available [9]. Sensor adherence was high, with 95% retention over three months and a median wear time of 89%, in a clinic where CGM had not previously been used.

The magnitude of the HbA1c reduction is consistent with, and at the upper end of, trials establishing CGM efficacy in high-income settings. DIAMOND reported 1.0% (95% CI 0.5–1.5) over 24 weeks in adults on multiple daily injections, and CITY 0.5% (95% CI 0.2–0.8) with a 10-percentage point TIR gain in 14-to 24-year-olds, with consistent findings elsewhere [2, 3, 4, 14]. Baseline HbA1c in our cohort far exceeded the 8.5–9.5% of those trials, and greater room for improvement probably explains part of the difference. The closest published comparison, McClure Yauch and colleagues’ East African study, described glycemic burden cross-sectionally, reporting a mean TIR of about 37% and TAR above 50%, close to our cohort at enrolment, but did not measure within-person change. Baker and colleagues in Rwanda and Gomber and colleagues in Malawi described feasibility and early improvement without pre-post HbA1c as an endpoint [6, 7, 8]. This study adds prospective pre-post HbA1c, biweekly CGM profiles and quantified sensor adherence from a Kenyan public-sector clinic.

Baseline characteristics reflect wider constraints on regional T1D care. Most participants (82.5%) used short-acting rather than rapid-acting insulin, and improvement on these regimens suggests that better glucose visibility yields benefit even where pharmacological options are limited [10]. The baseline DKA burden, with 42.5% reporting at least one episode in the preceding year, is consistent with regional data [12].

The reduction in diabetes-related admissions, from 7 of 40 to 1 of 38, was predominantly a reduction in hyperglycemic and ketotic presentations, which is directionally consistent with the fall in TAR and in the fraction of values above 13.9 mmol/L, and with population data showing that CGM adoption lowers both ketoacidosis and severe hypoglycemia, most markedly at HbA1c of 10% or above [13, 15, 16, 17]. With one event in the follow-up period, however, this observation is descriptive only.

### Limitations

The central limitation is that this was an uncontrolled study of a combined intervention. Participants received CGM together with structured education and systematic insulin titration, and with a modest increase in the frequency of clinical contact relative to the six-weekly review that constitutes standard management at this clinic. The observed change, therefore, may nobe attributed to CGM alone. It is relevant, however, that the increment in visit frequency was small: participants attended approximately two more contacts over three months than routine care would have provided, which makes clinical attention and contact frequency a less plausible sole explanation than it would be against a background of less frequent review. Structured education and systematic titration remain plausible contributors. Because no randomized comparison is planned in Kenya, separating the components of the package will require either a future trial or systematic outcome monitoring within a phased program, and in the interim, the package, rather than CGM in isolation, is what the data explains.

Regression to the mean is a second and independent explanation that this design cannot exclude. Participants entered with a high mean HbA1c of 10.82%, and cohorts selected at a high baseline value tend to move towards the population mean on repeat measurement irrespective of any intervention. Pre-enrolment HbA1c trajectories were not collected, as serial historical values were outside the scope of the parent study, so we cannot demonstrate that the observed change departs from a pre-existing trend. Two observations bear on this without resolving it: enrolment HbA1c spanned 6.50% to 15.30%, so the cohort was not selected on a high threshold value, and the direction and magnitude of change are consistent with randomized trials in which regression to the mean is controlled by design [2, 3, 4]. Participation itself may also have altered behavior independently of the glucose data, and the study cannot distinguish this effect.

Further limitations follow from the design. The sample of 40 was set by the parent usability study rather than by a power calculation, and it limits the precision of every estimate and precludes subgroup analysis or inference about admissions. Enrolment from a single tertiary center of participants in active follow-up, who were able to attend five visits over three months, selects for engagement and limits generalizability to district-level services and to patients who attend irregularly. Three months matches the duration of landmark trials and the period of peak engagement [18], but says nothing about durability once sensor access ends, which is the central question where CGM is unfunded [2, 3, 9, 10]. Pre-enrolment admissions and hypoglycemia frequency were ascertained by recall, while follow-up events were captured prospectively. Finally, hemoglobinopathy and anemia status were not systematically ascertained, and both can affect the interpretation of HbA1c in this population.

### Implications for policy in Kenya

The question facing Kenyan policymakers is not whether CGM works. That has been settled by randomized trials and meta-analyses, and no Kenyan trial is planned or needed to re-establish it [2, 3, 4, 14, 15]. The open question is whether an intervention proven in well-resourced systems retains its benefit, and can actually be delivered, under the conditions of a Kenyan public clinic.

Three findings are directly relevant to such a decision. First, in a clinic with no prior CGM experience, 95% of participants completed three months and median wear time was 89%, with adherence sustained in a population in which most families fall in the lower income strata and most participants are of school age. Second, the direction and magnitude of glycemic change are consistent with the randomized literature and were achieved despite human rather than analogue insulin in 82.5% of participants, which suggests that benefit is not contingent on first having modernized insulin supply. Third, safety was maintained, with time below 3.0 mmol/L unchanged and a single hypoglycemia of admission, in a setting where glucagon is rarely available.

A mean HbA1c above 10.8%, TIR far below target, and 42.5% of participants reporting at least one episode of ketoacidosis in the preceding year describe the standard of control that current practice produces.

## Conclusions

In this single-arm pilot at a national referral hospital in Kenya, three months of CGM use, supported by structured education and systematic insulin titration, was associated with a reduction in HbA1c and with progressive improvement in CGM-derived glycemic metrics among children, adolescents and young adults with T1D, alongside high sensor adherence and without an increase in clinically significant hypoglycemia. Because the design was uncontrolled and CGM was delivered as part of a package, the effect estimate is not unbiased. They support inclusion of CGM in national guidance for a defined high-risk group, delivered with the education and titration package alongside which it was tested, and coupled to routine outcome monitoring and formal costing so that durability and cost-effectiveness are established as the program proceeds.

## Data Availability

All data produced in the present work are contained in the manuscript

## Author Contributions

The authors confirm contribution to the paper as follows: study conception and design: Marbán-Castro E, Kamau Y; development and piloting of study instruments: Mungai L, Amolo P, Karume AK, Singh P; data collection: Mungai L, Amolo P, Karume AK, Munene-Waweru P, Nailaen C, Mule P; technical support during data collection: Marbán-Castro E, Kamau Y; data management and statistical analysis: Botella N; analysis and interpretation of results: Mungai L, Amolo P, Karume AK, Botella N, Marbán-Castro E, Kamau Y, Singh P; draft manuscript preparation: Mungai L. All authors reviewed the results and approved the final version of the manuscript.

## Acknowledgments

We would like to thank the guardians and all participants in this study

## Data Availability

The data underlying the findings reported in this article are not publicly available because they contain information that could compromise the privacy of participating children. De-identified data supporting the conclusions may be requested from Kenyatta National Hospital-University of Nairobi Ethics Review Committee at and will be released subject to approval by the Research Committee and completion of a data transfer agreement.

## Related Publication

The qualitative acceptability and feasibility findings from the same cohort are reported separately [22]. The present manuscript reports the clinical and glycemic outcomes only, and there is no overlap of results between the two reports.

## Electronic Supplementary Information

Supplementary Table S1, the completed TREND checklist for this study, is available as a separate file.

## References

1. Diabetes Control and Complications Trial Research Group. 1993. The effect of intensive treatment of diabetes on the development and progression of long-term complications in insulin-dependent diabetes mellitus. New England Journal of Medicine 329:977–86 10.1056/NEJM199309303291401

2. Beck RW, Riddlesworth T, Ruedy K, Ahmann A, Bergenstal R, et al. 2017. Effect of continuous glucose monitoring on glycemic control in adults with type 1 diabetes using insulin injections: the DIAMOND randomized clinical trial. Journal of the American Medical Association 317:371–78 10.1001/jama.2016.19975

3. Laffel LM, Kanapka LG, Beck RW, Bergamo K, Clements MA, et al. 2020. Effect of continuous glucose monitoring on glycemic control in adolescents and young adults with type 1 diabetes: the CITY randomized clinical trial. Journal of the American Medical Association 323:2388–96 10.1001/jama.2020.6940

4. Teo E, Hassan N, Tam W, Koh S. 2022. Effectiveness of continuous glucose monitoring in maintaining glycaemic control among people with type 1 diabetes mellitus: a systematic review of randomised controlled trials and meta-analysis. Diabetologia 65:604–19 10.1007/s00125-021-05648-4

5. Battelino T, Danne T, Bergenstal RM, Amiel SA, Beck R, et al. 2019. Clinical targets for continuous glucose monitoring data interpretation: recommendations from the international consensus on time in range. Diabetes Care 42:1593–1603 10.2337/dci19-0028

6. McClure Yauch L, Velazquez E, Piloya-Were T, Wainaina Mungai L, Omar A, et al. 2020. Continuous glucose monitoring assessment of metabolic control in East African children and young adults with type 1 diabetes: a pilot and feasibility study. Endocrinology, Diabetes & Metabolism 3:e00135 10.1002/edm2.135

7. Baker J, Cappon G, Habineza JC, Basch CH, Mey S, et al. 2025. Continuous glucose monitoring among patients with type 1 diabetes in Rwanda (CAPT1D) phase I: prospective observational feasibility study. JMIR Formative Research 9:e64585 10.2196/64585

8. Gomber A, Valeta F, Coates MM, Trujillo C, Ferrari G, et al. 2024. Feasibility of continuous glucose monitoring in patients with type 1 diabetes at two district hospitals in Neno, Malawi: a randomised controlled trial. BMJ Open 14:e075554 10.1136/bmjopen-2023-075554

9. Ogle GD, Kim H, Middlehurst AC, Silink M, Jenkins AJ. 2016. Financial costs for families of children with type 1 diabetes in lower-income countries. Diabetic Medicine 33:820–26 10.1111/dme.12997

10. Bahendeka S, Ramaiya K, Swai AB, Otieno F, Bajaj S, et al. 2018. EADSG guidelines: insulin therapy in diabetes. Diabetes Therapy 9:449–92 10.1007/s13300-018-0384-6

11. Hall V, Thomsen RW, Henriksen O, Lohse N. 2011. Diabetes in sub-Saharan Africa 1999–2011: epidemiology and public health implications. A systematic review. BMC Public Health 11:564 10.1186/1471-2458-11-564

12. Cassambai S, Godia P, Bahendeka S, Omodara D, Ikhile D, et al. 2026. Strengthening diabetes care in sub-Saharan Africa: health systems, digital innovations, lifestyle interventions, and socioeconomic dimensions. The Lancet Diabetes & Endocrinology 14:523–32 10.1016/S2213-8587(26)00035-5

13. Maahs DM, Hermann JM, Holman N, Foster NC, Kapellen TM, et al. 2015. Rates of diabetic ketoacidosis: international comparison with 49,859 pediatric patients with type 1 diabetes from England, Wales, the U.S., Austria, and Germany. Diabetes Care 38:1876–82 10.2337/dc15-0780

14. Juvenile Diabetes Research Foundation Continuous Glucose Monitoring Study Group. 2008. Continuous glucose monitoring and intensive treatment of type 1 diabetes. New England Journal of Medicine 359:1464–76 10.1056/NEJMoa0805017

15. Karges B, Tittel SR, Bey A, Freiberg C, Klinkert C, et al. 2023. Continuous glucose monitoring versus blood glucose monitoring for risk of severe hypoglycaemia and diabetic ketoacidosis in children, adolescents, and young adults with type 1 diabetes: a population-based study. The Lancet Diabetes & Endocrinology 11:314–23 10.1016/S2213-8587(23)00061-X

16. Rodríguez de Vera Gómez P, Rodríguez Jiménez B, Mayoral Sánchez E, Jódar Sánchez F, Ravé García R, et al. 2025. Intermittently scanned continuous glucose monitoring adoption decreases diabetic ketoacidosis hospitalizations and healthcare costs in adults with type 1 diabetes. Diabetes Research and Clinical Practice 226:112372 10.1016/j.diabres.2025.112372

17. Mustonen J, Rautiainen P, Lamidi ML, Lavikainen P, Martikainen J, et al. 2023. The use of isCGM leads to marked reduction in severe hypoglycemia requiring emergency medical service or hospital admission and diabetic ketoacidosis in adult type 1 diabetes patients. Acta Diabetologica 60:891–98 10.1007/s00592-023-02079-y

18. Jensen MH, Cichosz SL, Gustenhoff P, Nikontovic A, Hejlesen O, et al. 2022. Long-term glucose-lowering effect of intermittently scanned continuous glucose monitoring for type 1 diabetes patients in poor glycaemic control from Region North Denmark: an observational real-world cohort study. PLOS One 17:e0274626 10.1371/journal.pone.0274626

19. International Diabetes Federation. 2025. IDF Diabetes Atlas. 11th Edition. Brussels: International Diabetes Federation

20. Diabetes Control and Complications Trial/Epidemiology of Diabetes Interventions and Complications Study Research Group. 2016. Intensive diabetes treatment and cardiovascular outcomes in type 1 diabetes: the DCCT/EDIC study 30-year follow-up. Diabetes Care 39:686–93 10.2337/dc15-1990

21. Marbán-Castro E, Muhwava L, Kamau Y, Safary E, Rheeder P, et al. 2024. Implementation research: a protocol for two three-arm pragmatic randomised controlled trials on continuous glucose monitoring devices in people with type 1 diabetes in South Africa and Kenya. Trials 25:331 10.1186/s13063-024-08132-7

22. Karume AK, Amolo P, Mungai L, Moraa H, Arunga T, et al. 2026. Acceptability, feasibility, and user experiences of continuous glucose monitoring in type 1 diabetes in Kenya: perspectives of adolescents, young adults, caregivers, and healthcare providers. medRxiv, preprint posted 18 August 2026 10.64898/2026.08.16.26359665

23. Des Jarlais DC, Lyles C, Crepaz N, and the TREND Group. 2004. Improving the reporting quality of nonrandomized evaluations of behavioral and public health interventions: the TREND statement. American Journal of Public Health 94:361–66 10.2105/ajph.94.3.361

24. Battelino T, Alexander CM, Amiel SA, Arreaza-Rubin G, Beck RW, et al. 2023. Continuous glucose monitoring and metrics for clinical trials: an international consensus statement. The Lancet Diabetes & Endocrinology 11:42–57 10.1016/S2213-8587(22)00319-9

25. Ngwiri T, Were F, Predieri B, Ngugi P, Iughetti L. 2015. Glycemic control in Kenyan children and adolescents with type 1 diabetes mellitus. International Journal of Endocrinology 2015:761759 10.1155/2015/761759

